# Independent Evaluation of Deep Learning Models for Detecting Focal Cortical Dysplasia

**DOI:** 10.1101/2025.04.06.25325315

**Authors:** Helene Kaas, Martin Prener, Melanie Ganz, Gitte M Knudsen, Lars H Pinborg, Vincent Beliveau

## Abstract

**Objective:** The objective of this study is to perform an independent assessment of the diagnostic utility of three state-of-the-art tools for the detection of focal cortical dysplasia (FCD) from Magnetic Resonance images (MRI). These tools include DeepFCD, the Multi-center Epilepsy Lesion Detection (MELD) Classifier, and the MELDGraph.

**Methods:** T1-weighted and fluid-attenuated inversion recovery MR images from 101 epilepsy patients with FCD and 101 age- and sex-matched epilepsy patients without FCD were included. Classifiers were evaluated at a patient-level by their ability to correctly identify the presence of any FCD lesions, and at a lesion-level by their capacity to identify lesions within the region delineated by the neuroradiologist in the MRI report. A calibrated threshold for DeepFCD prediction probabilities was empirically determined to improve classifier specificity. Test-retest consistency of the classifiers was measured using the Dice coefficient on repeated MRI scans of 21 individuals.

**Results:** For assessments at patient-level, high false positive rates were prominent, with the MELDClassifier achieving 52% accuracy (sensitivity=91%, specificity=14%). MELDGraph performed with accuracy up to 61% (sensitivity=76%, specificity=47%) and DeepFCD reached 56% accuracy (sensitivity=62%, specificity=50%) at an empirically determined threshold of 0.90. When investigating specific lesions, the MELDClassifier performed with a sensitivity of 91% and positive predictive value (PPV) of 13%, and MELDGraph performed with a sensitivity of 69% and PPV of 36%, whereas the DeepFCD achieved a sensitivity of 100% and PPV of 4%. Test-retest reliability was low, with an average [min, max] Dice coefficient of 0.28 [0.0, 1.0] for MELDClassifier, 0.38 [0.0, 1.0] for MELDGraph with harmonization and 0.35 [0.05, 0.54] for DeepFCD.

**Significance:** This study highlights the current limitations of using deep learning models in FCD diagnosis and emphasizes the need to enhance the tools’ accuracy, reliability, and interpretability to improve their clinical utility in epilepsy diagnosis.

**Key points:**

- State-of-the-art deep learning tools for identifying focal cortical dysplasia perform with high sensitivity ranging from 69% to 100%
- When predicting the presence of any lesion within epilepsy patients’ MRI scans, the classifiers performed with accuracies ranging from 52% to 61%
- The average false positive count per patient ranged from 0.49 ± 0.66 (MELDGraph) to 32.71 ± 14.35 (DeepFCD).
- All three classifiers had low test-retest consistency, suggesting that the predictions may be strongly influenced by the noise in the images.

## Introduction

Focal cortical dysplasia (FCD) is one of the leading causes of drug-resistant epilepsy^1^. The presentation of FCD on Magnetic Resonance Imaging (MRI) often involves a combination of imaging findings, each of which is subtle on its own (Figure 1) and is often missed, leading to ‘MRI-negative’ misclassifications^2^. This can delay or prevent diagnosis and later referral for epilepsy surgery or lead to less accurate pre-surgical diagnoses, resulting in suboptimal outcomes^3,4^. According to the International League Against Epilepsy, clinical and histological features of FCD are categorized into subtypes I-III^5^. Common radiological features, include (1) increased cortical thickness, (2) blurring of the gray/white matter junction, (3) transmantle sign, (4) abnormal gyral/sulcal pattern, and (5) Fluid attenuated inversion recovery (FLAIR) hyperintensity^6,2^. The prevalence of these characteristics depends on the clinicopathological subtype^7,8^.

**Figure 1.**
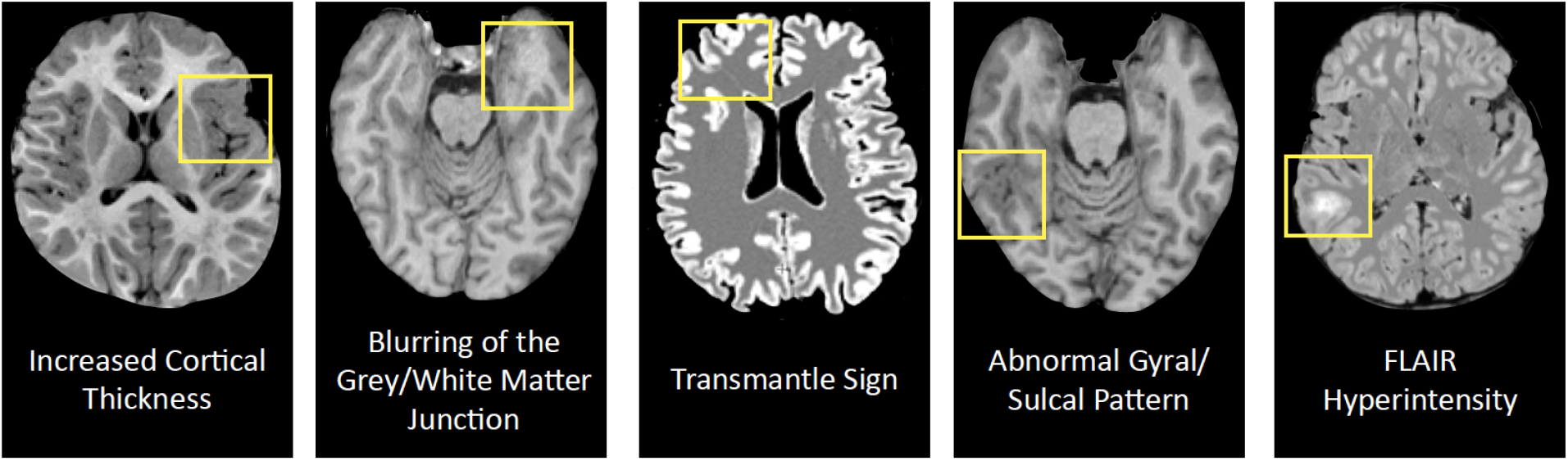
Common radiological features of Focal Cortical Dysplasia (FCD) in Magnetic Resonance Imaging (MRI).

Considerable efforts have been made to improve the detection of FCD from MR images. By leveraging technological advancements in image processing brought forward by deep learning, multiple models have been proposed to further improve the state-of-the-art of FCD detection^9,10,11,12,13^. To date, the three most promising tools for automatic FCD detection are the Multi-center Epilepsy Lesion Detection (MELD) Classifier^14^, MELDGraph^15,16^, and DeepFCD^17^. Validation performed by the authors of these models on independent datasets has demonstrated sensitivities of 88% (MELDClassifier), 72% (MELDGraph), and 83% (DeepFCD), and specificities of 17% (MELDClassifier), 56% (MELDGraph), and 89% (DeepFCD)^14,16,17,15^. Although promising, these results derive from comparisons of epilepsy patients with healthy controls with no additional brain abnormalities. Therefore, these reported performances are potentially overly optimistic as they do not accurately represent the clinical settings where the tools would be solely applied to patients suspected of epilepsy who may indeed present a variety of other brain abnormalities.

This study compares the performance of the state-of-the-art FCD classifiers against a ground truth established in the clinical setting by neuroradiologists in a large cohort of epilepsy patients with and without FCD. An evaluation of the tools, entirely independent of the developers and the data source, is critical for establishing their usefulness as a clinical diagnostic tool.

## Materials and Methods

Performance evaluations were conducted on three different state-of-the-art classifiers, all consisting of a processing pipeline that extracts data from T1-weighted (T1w) and FLAIR MRI scans to make lesion-level predictions. An overview of the workflow for each of the three models is presented in Figure 2.

**Figure 2.**
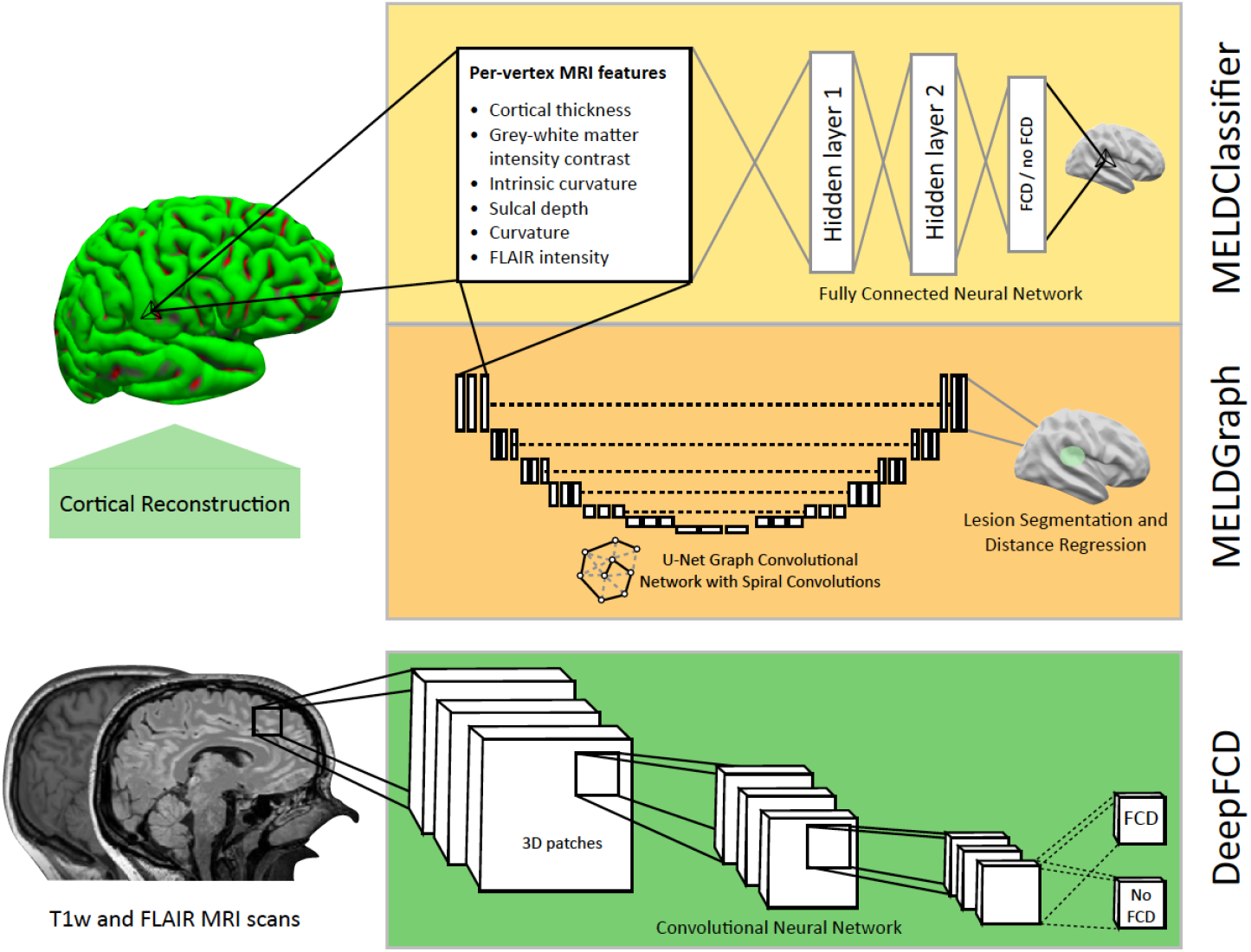
Overview of the classifier workflow for MELDClassifier, MELDGraph, and DeepFCD. All classifiers utilize both a T1 weighted (T1w) and a fluid-attenuated inversion recovery sequence (FLAIR). Cortical reconstruction is achieved through Freesurfer’s *recon-all* function which is utilized by MELDClassifier and MELDGraph to make per-vertex predictions, whereas the DeepFCD tool makes voxel-based predictions by extracting 3D patches directly from the MRI volumes.

### MELDClassifier

The MELDClassifier extracts cortical features from MRI scans to make per-vertex predictions of FCD presence using a fully connected neural network. The MRI-derived input features include measures of cortical thickness, gray-white matter contrast, and curvature. These features are extracted using FreeSurfer’s *recon-all* software^18^. The classifier uses ComBat to harmonize MRI data by adjusting for scanner- and sequence-specific site effects while preserving biological variation linked to age, sex, and FCD status.

### MELDGraph

The MELDGraph is a graph convolutional network, in which the cortical surface is processed as a graph structure with vertices and edges representing nodes and spatial connections, respectively. The graph convolutional network incorporates spiral convulsions, distance loss, and spatial loss within a U-Net architecture to model spatial dependencies on the cortical surface. It uses the same FreeSurfer-extracted features and ComBat harmonization as MELDClassifier, and applies Monte Carlo dropout^19^ to estimate prediction confidence via averaged softmax probabilities.

### DeepFCD

The DeepFCD tool incorporates a convolutional neural network (CNN), to make per-voxel predictions of FCD in MRI scans. The CNN input of the DeepFCD tool is composed of 3D voxel patches of 16×16×16 voxels, extracted from scans preprocessed using DeepMask, an in-house skull-stripping method. The network applies the Monte Carlo dropout method^19^ to simulate a Bayesian CNN. From these distributions, prediction probabilities are calculated as the mean, while prediction uncertainty is defined as the inverse of the variance. Median uncertainties across voxels in a predicted FCD lesion are extracted and normalized between 0 and 1 to obtain measures of confidence. Confidence is thus a relative term restricted to predictions within a single patient. Prediction probabilities are not constrained by a fixed threshold, leading to high numbers of predictions per patient. An empirically determined threshold is therefore required to increase specificity.

### Data Extraction and Selection of Patients and MRI Sequences

Patients with ICD-10 code G40* were identified from Copenhagen University Hospital Rigshospitalet (RH), all of whom had a contact in the Neuroscience Center of RH between 2017 and 2023. MR images and their corresponding report were retrieved in bulk from the medical image archiving and communication system of RH. Due to limitations imposed by the MELD tools, a unique T1w and FLAIR sequence pair had to be selected for analysis. The most numerous sequence pair was chosen to maximize participant inclusion. The MRI sequence parameters of the chosen sequences are described in Supplementary Text 1. Only patients with the required sequences acquired on 3T scanners were included. Patients were excluded if they were under the age of 3, or if the relevant MRI scans were only available after intracranial surgery or other interventions, that could cause structural changes to the brain. Patients with radiology reports describing the presence or absence of FCD were first identified using regular expressions. Then, to confirm the classification of FCD-positive patients, a manual assessment of patients’ clinical notes was performed. This assessment relied on radiologists’ MRI descriptions. All MRI scans were interpreted by two neuroradiologists who are members of the Danish Epilepsy Surgery Group and thus subspecialized in MRI diagnostics in epilepsy. In case of high uncertainty, the assessment was supported by congruence between MRI-identified lesions and other diagnostic modalities such as EEG.

A total of 121 FCD-positive patients were included and paired with an FCD-negative patient using the Nearest Neighbour distance matching method, based on age and sex differences. Clinical notes of selected FCD-negative patients were manually reviewed to confirm their epilepsy diagnosis. Of these, 101 FCD-positive patients and their FCD-negative matches were used to calculate classifier performance metrics, while the remaining 20 pairs were independently set aside for classifier calibration. Classifier accuracy with the 95% confidence interval as a function of the number of sampled patients is presented in Supplementary Figure 2.

### Data Processing

Cortical reconstruction was performed on the selected scans for each subject using FreeSurfer v7.2.0 and the *recon-all* pipeline. Quality control was ensured via the Deep-MI/fsqc^20^ protocol. A new site was obtained from the MELD consortium and site-specific harmonization parameters were computed using the independent balanced cohort of 20 FCD-positive and -negative pairs.

The independent cohort was also utilized to calibrate an empirical threshold for the DeepFCD tool’s prediction probability score by determining the maximal value of Youden’s index^21^. A single threshold was determined for both patient- and lesion-level assessments, based on patient-level definitions of true and false predictions, as a negative prediction could not be clearly defined at the lesion-level.

### Evaluation Metrics

Classifiers’ performance was evaluated by comparing predictions with neuroradiologists’ assessments. Specificity, sensitivity, accuracy, positive predictive value (PPV), negative predictive value (NPV), and average false positive (FP) per patient were used as evaluation metrics. The 95% confidence intervals for evaluation metrics were determined using a nonparametric bootstrap of 10,000 iterations and resampling with replacement. The same method was used to compare performance metrics across classifiers. Outputs from DeepFCD were analyzed with two thresholds: (1) a fixed prediction probability threshold of 0.7, adopted from a previous validation of the classifier^17^, and (2) an empirically determined threshold (see Data Processing).

At the patient-level, tools were assessed for their ability to diagnose FCD using a binary approach, describing the presence or absence of predictions per patient, regardless of lesion localization. Predictions at this level are described as key for FCD detection^2^, since a tool that provides a clear diagnostic outcome is more likely to assist clinicians in refining their hypotheses and further planning. Performance was assessed on a lesion-level, in which predicted clusters were considered true positive (TP) when overlapping with the neuroradiologist-defined region. At the lesion-level, all available MRI descriptions provided by neuroradiologists were assessed in concurrency with seizure freedom, post-operative MRI scans, and histology reports in cases involving surgery. The information was used to identify the exact locations of FCD-lesions, often on a gyral/sulcal level, as presented in Figure 3. Superimposition of the Desikan-Killiany brain atlas^22^ with native MRI FCD-positive scans as well as available post-surgical scans (n=32) enabled precise localization of FCD regions. Since there is no clear criteria defining negative lesion-level predictions, metrics relying on true negatives (TN) (i.e., accuracy, specificity, and NPV) were not calculated at this level. Lesions within major visible structural pathologies were disregarded from the evaluation.

**Figure 3.**
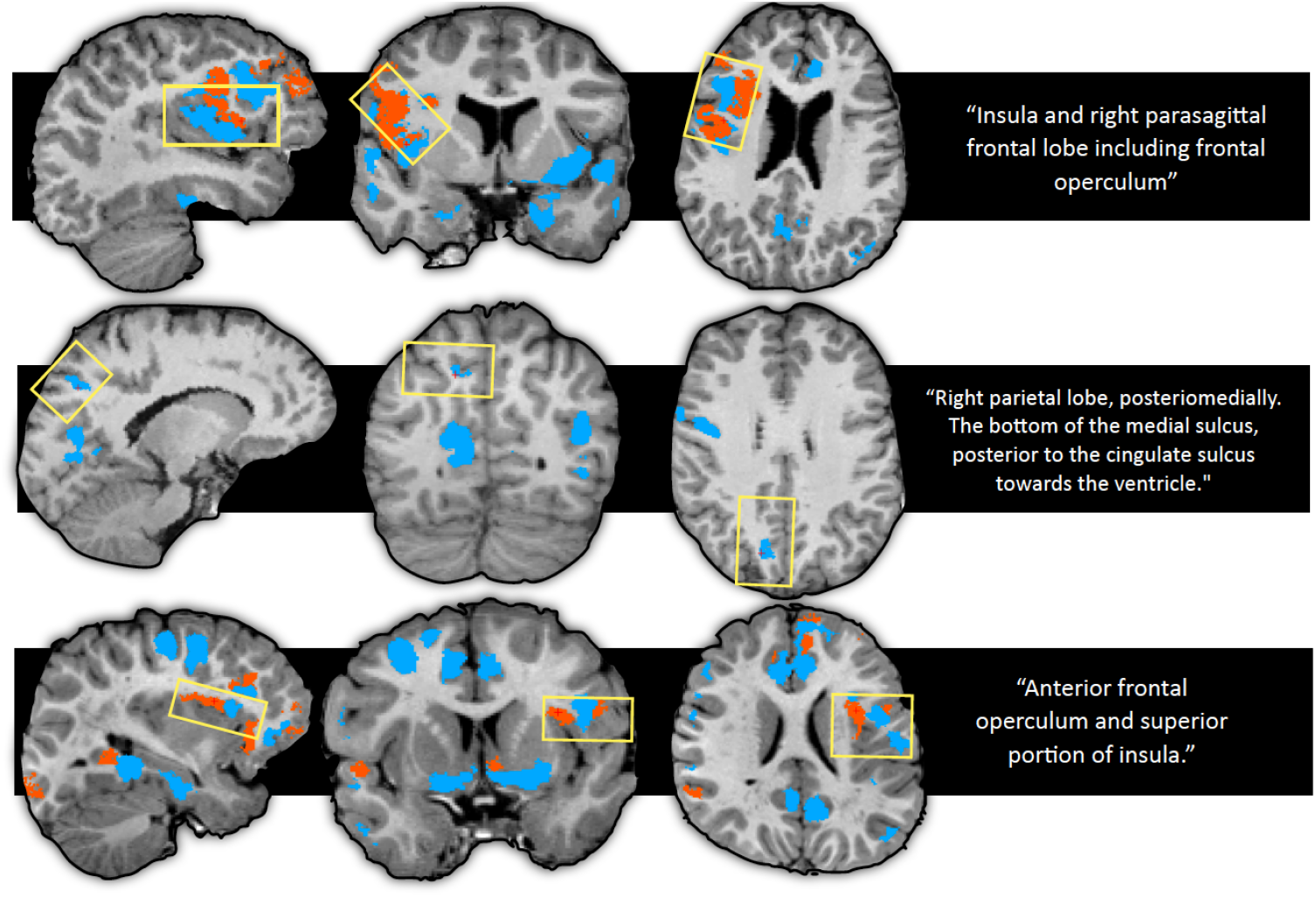
Overview of lesion-level evaluation. Ground truths were established based on neuroradiologists’ descriptions of lesion location, often on a gyral/sulcal level, obtained from MRI reports in congruence with postoperative scans (if available) and information about seizure freedom. This enabled the precise localisation of FCD regions (yellow boxes) and true positives (TPs) within the prediction maps of the classifiers (blue and orange cluster).

### Test-Retest Reliability

In 21 patients, two temporally separate consecutive MRI scans were performed. Test-retest reliability was evaluated by comparing the overlap of predicted lesion maps from two MRIs of the same individuals using the Dice coefficient. Scans were aligned using rigid registration. Notable observations regarding the MELDClassifier’s and MELDGraph’s performances were cases where the classifiers consistently did not produce any predictions in both MRIs for the same patient. These occurrences were interpreted as perfect agreements, thus attributing Dice scores of one.

### DeepFCD and MELDGraph Prediction Distributions

Distributions of confidence values for DeepFCD and MELDGraph were visualized using density plots. The association between DeepFCD prediction rank and probability was assessed using Spearman’s correlation. Furthermore, the distributional difference between DeepFCD predicted probabilities of FP and TP predictions was tested using the Wilcoxon signed-rank test, comparing aggregated prediction probabilities across subjects.

### Influencing Factors on Classifier Performance

To investigate factors that may influence the performance of the classifiers, additional tests were performed. Dice coefficients between predicted lesion maps of MELDGraph with and without harmonization were calculated to evaluate its influence.

Bootstrapping of 10,000 iterations and resampling with replacement was used to assess differences in sensitivity between FCD-positive patients and several subgroups, including patients who underwent epilepsy surgery (n=47), those achieving seizure freedom post-surgery (n=29), and patients initially considered MRI-negative because their first MRI reports did not mention FCD, but later MRI- or histology reports revealed an FCD lesion (n=49). For each comparison, respective subgroups were subtracted from the remaining FCD-positive cohort to assume independence.

Similarly, bootstrapping was used to evaluate the average FP count per patient for scans with minor image quality artifacts (n=117), patients with major structural brain pathology, such as schizencephaly, ischemia, cerebral atrophy, or large arachnoid cysts, resulting in widespread morphological changes at the lobar or hemispheric level (n=7), or patients only ever considered MRI-negative (n=33) in comparison to the remaining full cohort. For each comparison, subgroups were subtracted from the full cohort to assume independence.

Spearman’s correlation coefficient was utilized to examine relations between patient age and FP count per patient. Predictions overlapping with regions of cortical pathology other than FCD were noted and compared to the total FP count per classifier. Classifier performance was described among patients with two separate regions with FCD (n=6).

## Results

### Demographics

The balanced cohort included 101 FCD-positive patients and 101 FCD-negative epilepsy patients. Sixteen patients had histologically proven FCD, and 49 patients had FCD which had been overlooked in the initial radiological assessment but were identified later. Seven patients had larger structural pathologies in their MRI, of which two were FCD-positive. Thirteen patients were under the age of six. Table 1 summarizes the demographic information and lesion characteristics, with FCD subtypes classified according to International League Against Epilepsy criteria^5^. Distributions of patient age at scan, comparing patients with and without FCD is visualized in Supplementary Figure 1.

**Table 1:** Overview of demographic information and clinical features of patient cohorts.

| MR description | Participants | Sex [female:male] | Average age at scan (IQR) | Seizure debut (IQR) | Hemisphere (%) | Surgery (%) | Pathology (%) |
| --- | --- | --- | --- | --- | --- | --- | --- |
| **FCD-positive** | 101 | 48:53 | 24.5 (3-70) | 11.7 (0-53) | Right (47.5%)<br>Left (52.5%)<br>Bilateral (0.1%) | Resection (42.0%)<br>LITT (5.0%)<br>Seizure-freedom:<br>2y FU: 23/38,<br>1y FU: 6/9,<br><1y FU: 1 | Histology confirmed FCD (n=42):<br>FCD type 2A (26.2%)<br>FCD type 2B (42.9%)<br>FCD type 1B (2.9%)<br>FCD type 3B (2.9%)<br>FCD type 3C (2.9%)<br>FCD, unspecified (19.0%) |
| **FCD-negative** | 101 | 48:53 | 24.5 (3-68) | 13.3 (0-65) | - | - | MRI negative (32.7%)<br>Other pathologies than FCD (67.3%):<br>Folding abnormalities, incl. polymicrogyria and migration, abnormalities,<br>Heterotopia,<br>Encephalocele,<br>Perinatal ischemia,<br>Infarction,<br>Trauma sequelae,<br>Tumor, incl. DNET, ganglioglioma, Hemorrhage, SWS and MTS |

*FCD* Focal Cortical Dysplasia, *MRI* Magnetic Resonance Imaging, *LITT* Laser Interstitial Thermal Therapy, *MTS* Mesial Temporal Sclerosis, *SWS* Sturge Weber Syndrome, *DNET* Dysembryoplastic Neuroepithelial Tumor, *y* year, *FU* Follow up

### Patient-level Classification

An overview of the total counts of TP, FP, TN, and false negative (FN), when compared to MRI reports by neuroradiologists, can be found in Supplementary Table 1. Classifier performance metrics, including accuracy, specificity, sensitivity, PPV, NPV, and FP count per patient are presented in Figure 4A and Supplementary Table 2.

**Figure 4.**
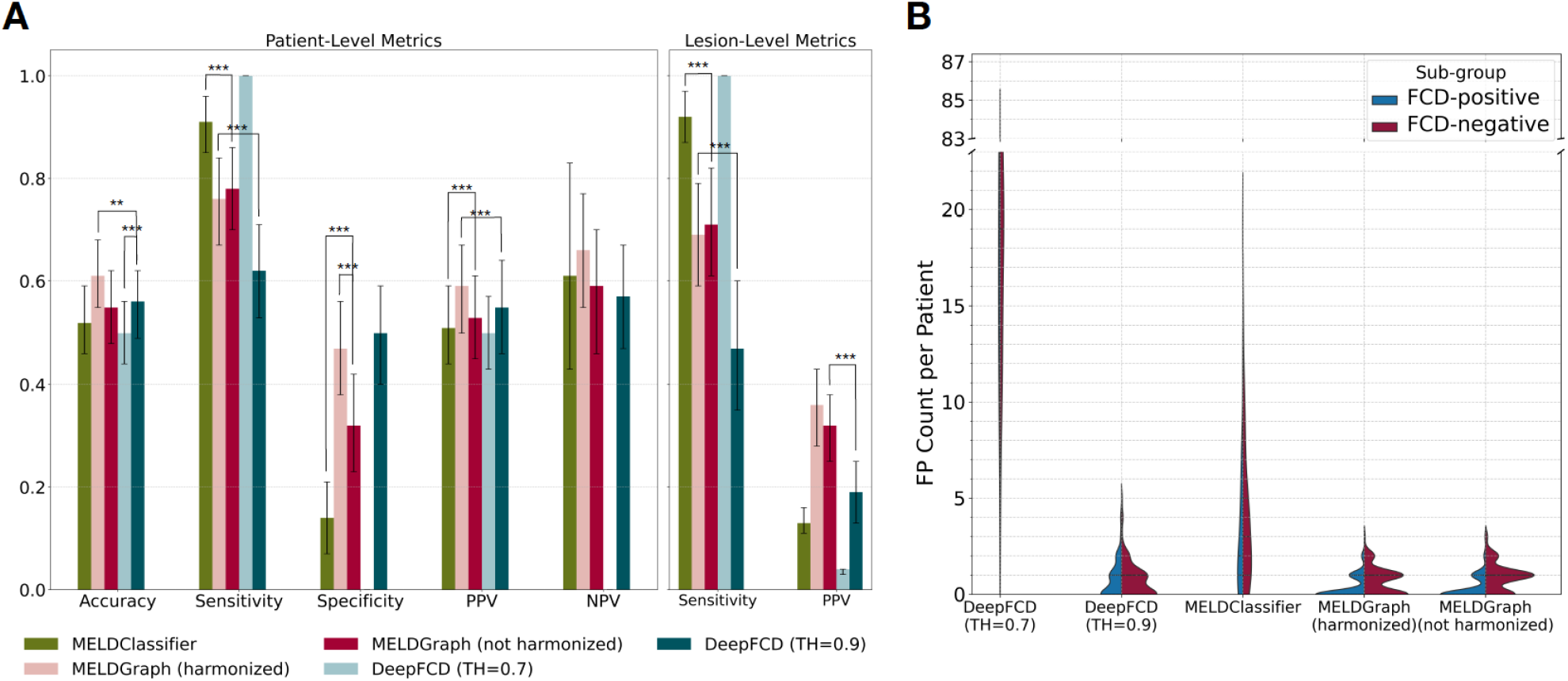
(A) Summary of Classifier Performance. The figure presents the accuracy, sensitivity, specificity, positive predictive value (PPV) and negative predictive value (NPV) for the MELDClassifier, MELDGraph, and DeepFCD at patient-as well as sensitivity and PPV at lesion-levels. Error bars indicate 95% confidence intervals and stars indicate statistical significance. (B) False positive (FP) count per patient for the MELDClassifier, MELDGraph and DeepFCD at lesion-level. DeepFCD output is held at a prediction probability threshold (TH) of 0.7, and at calibrated threshold of 0.90 (patient-level) derived from the ROC curve. Performance metrics for MELDGraph are presented with and without harmonisation.

The MELDClassifier identified the presence of any FCD lesions with an accuracy of 52% (sensitivity=91%, specificity=14%), PPV of 51%, and NPV of 61%. The MELDGraph performed with an accuracy of 61% (sensitivity=76%, specificity=47%) PPV of 59% and NPV of 66% with harmonization and 55% (sensitivity=78%, specificity=32%) PPV of 53% and NPV of 59% without harmonization.

The ROC curve for DeepFCD is presented in Figure 5C. The calibrated threshold for DeepFCD prediction probabilities was determined to be 0.90. When using the predefined probability threshold of 0.7, DeepFCD made predictions on all subjects, while its accuracy rose to 56% (sensitivity=62%, specificity=50%) with the calibrated threshold.

**Figure 5.**
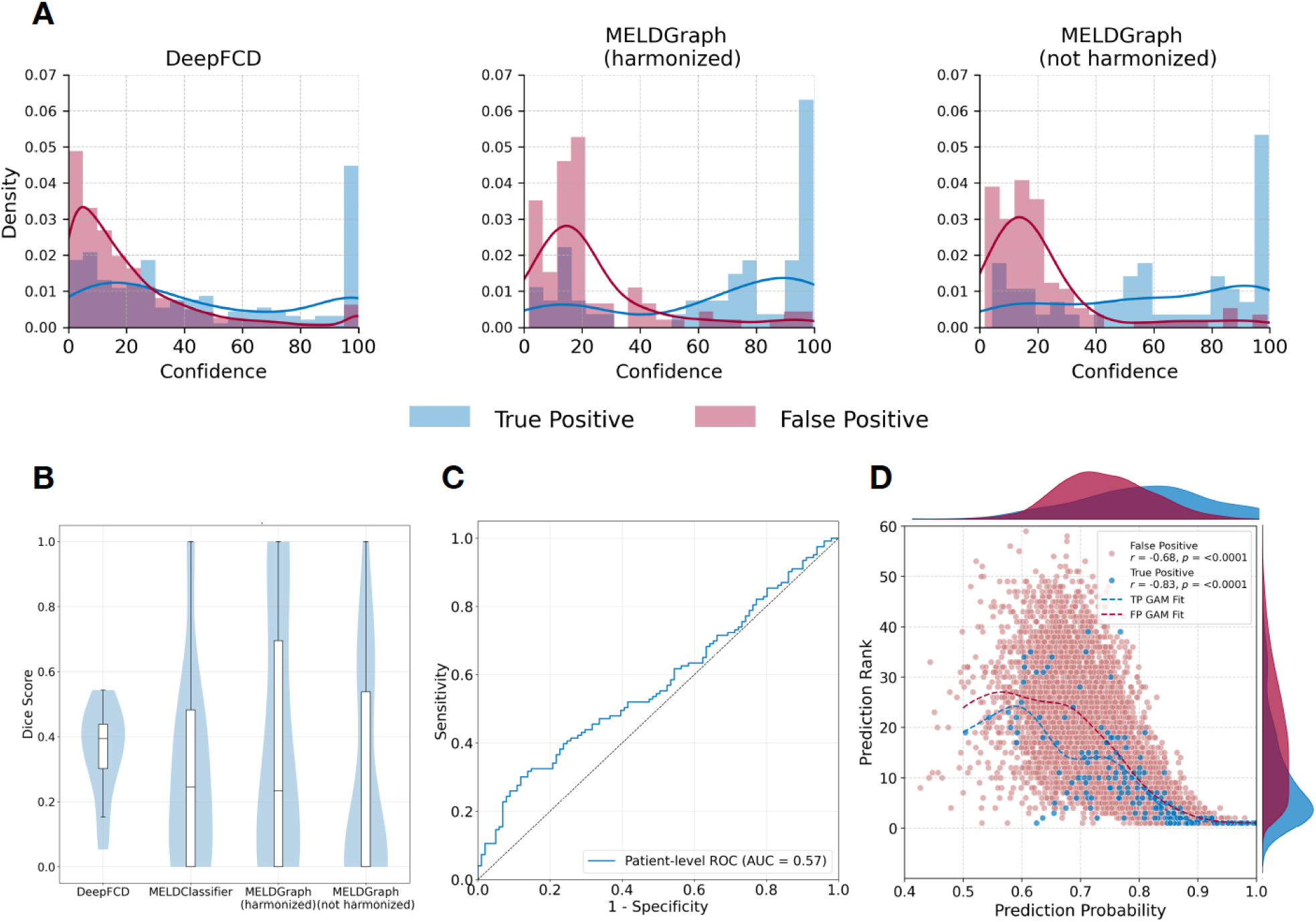
(A) Distribution of confidence values for DeepFCD, MELDGraph (harmonized), and MELDGraph (not harmonized). FP predictions largely center around lower values, while TP predictions are distributed bimodally in all three cases. (B) Distributions of Dice coefficients assessing the consistency of prediction outputs for the same subjects at two different time points (C) Receiving operating characteristic (ROC) curve for DeepFCD at patient-level. (D) Correlation and distributions of prediction probability values and ranking for DeepFCD. The correlation is represented by a generalized additive model (GAM). TP predictions have significantly higher ranking and probability values than FP predictions (p>0.001).

Accuracy and PPV improved significantly (p<0.001), when subjecting DeepFCD to an empirical threshold compared to the fixed threshold of 0.7. MELDGraph performed with significantly (p<0.01) better accuracy than all other classifiers and specificity and PPV improved significantly (p<0.001) with harmonization compared to without. Regardless of harmonization, MELDGraph showed significantly higher specificity and PPV (p<0.001) than MELDClassifier. However, MELDClassifier had significantly higher sensitivity (p<0.001) than both MELDGraph and DeepFCD thresholded at 0.9. Differences in specificity between MELDGraph and DeepFCD thresholded at 0.9 was insignificant.

### Lesion-level Classification

Neuroradiologist-confirmed lesions were identified by MELDClassifier with a sensitivity of 92% and PPV of 13%, whereas MELDGraph performed with a sensitivity of 69% and PPV of 36% with harmonization and a sensitivity of 71% and PPV of 32% without harmonization. DeepFCD performed with a sensitivity of 100% and PPV of 4% at the fixed threshold of 0.7 and with a sensitivity of 47% and PPV of 19% at the calibrated threshold of 0.9. The MELDClassifier produced an average of 3.51 ± 3.58 FP predictions per patient, compared to the MELDGraph which produced 0.49 ± 0.66 FP predictions with harmonisation and 0.59 ± 0.71 without harmonisation. DeepFCD produced an average of 32.71 ± 14.35 FP predictions per patient. Figure 4B presents the distributions of FP count per patient.

Regardless of harmonization, the MELDGraph performed with significantly (p<0.001) better PPV compared to the MELDClassifier and DeepFCD, whereas the MELDClassifier performed with significantly higher sensitivity than the MELDGraph and DeepFCD at a threshold of 0.9.

MELDGraph’s TP predictions had an average [min, max] confidence of 65.2 [1.7, 99.7]. TP predictions of DeepFCD had an average [min, max] confidence of 63.2 [0.4, 100] and a prediction probability of 0.80 [0.55, 0.87].

### Test-Retest Reliability

Dice coefficients were calculated to assess the consistency of prediction outputs for the same patient, when scanned at two different time points (mean interval [min–max]: 22.8 months [3.1–75.6]). The median [Q1, Q3] Dice coefficients across subjects were 0.23 [0.0, 0.70] for MELDGraph with harmonization and 0.0 [0.0, 0.54] without harmonization. For the MELDClassifier, the average Dice coefficient was 0.25 [0.0, 0.48], and for DeepFCD 0.39 [0.30, 0.44]. The distribution of Dice coefficients per classifier is presented in Figure 5B.

### DeepFCD and MELDGraph Prediction Distributions

The density distributions of prediction confidence values for DeepFCD and MELDGraph predictions are illustrated in Figure 5A. Notably, the confidence of FP outputs for both classifiers predominantly centered around lower values, indicating a general trend of low confidence in these predictions. In contrast, confidence distributions for TP outputs displayed a bimodal distribution with a peak near lower ranges and another close to 100. In 87.1% of cases, the TP were among the top five DeepFCD predictions with the highest confidence. Figure 5D shows the association between prediction probability and rank for DeepFCD. Spearman correlations between prediction probability and rank were significant for both FP (r(200)=−0.68, p<0.0001) and TP predictions (r(200)=−0.83, p<0.0001). Probability distributions were statistically analyzed using Wilcoxon signed-rank, which confirmed that the observed differences between FP and TP were significant (W=517.0, z=−6.62, p<0.0001). The median [Q1, Q3] aggregated FP probability was 0.73 [0.72, 0.74] and the median aggregated TP probability was 0.82 [0.74, 0.88].

### Influencing Factors on Classifier Performance

Dice coefficient was calculated to assess the similarity of the predictions from the MELDGraph classifier with and without harmonization. The results showed a median [Q1, Q3] Dice coefficient of 0.0 [0.0, 0.66] across subjects.

None of the classifiers showed significant differences in sensitivity between FCD-positive patients and subgroups, including patients who underwent epilepsy surgery, those achieving seizure freedom post-surgery, and patients initially considered MRI-negative, compared to the remaining FCD-positive cohort. The DeepFCD confidence scores for TP predictions were significantly lower (p=0.046) for the FCD-positive, MRI-negative subgroup compared to the remaining FCD-positive cohort.

Patients with minor artifacts in their MRI scans had significantly higher average FP counts per patient with the MELDClassifier (p<0.001). DeepFCD probability scores were significantly higher (p=0.021) for this subgroup, whereas no significant difference was found for DeepFCD or MELDGraph confidence scores.

Patients with major structural brain pathologies in their MRI scans had a significantly higher average FP count per patient for MELDGraph with (p<0.001) and without (p=0.010) harmonization. DeepFCD confidence scores were significantly higher (p<0.001) for this subgroup, whereas MELDGraph confidence scores showed no significant difference.

FCD-negative MRI-negative patients had a significantly higher average FP count per patient when predictions were made with MELDGraph without harmonization (p<0.024). DeepFCD confidence scores were significantly higher (p<0.001) for this subgroup, whereas MELDGraph confidence scores showed no significant difference. Patient age was positively associated with higher FP counts per patient for both DeepFCD, MELDClassifier, and MELDGraph with harmonization (DeepFCD: r(200)=−0.43, p>0.001, MELDClassifier: r(200)=−0.51, p>0.001, MELDGraph (harmonized): r(200)=−0.21, p=0.033, MELDGraph (not harmonized): r(200)=−0.03, p=0.721).

A total of 167 regions with cortical pathologies unrelated to FCD were identified within the full cohort. DeepFCD mistakenly identified 130 of these regions as FCDs, accounting for 4.2% of the total FPs. The MELDClassifier made predictions in 55 regions (7.8% of total FPs), while MELDGraph identified 27 of these regions with harmonization (27.6% of total FPs) and 39 without harmonization (32.8% of total FPs). Detected non-FCD cortical pathologies per classifier can be found in Supplementary Table 3.

In patients with two distinct FCD regions, DeepFCD identified both regions in all patients. MELDClassifier identified both regions in four out of six patients and made at least one correct prediction in five patients. Both with and without harmonization, MELDGraph only identified both FCD regions in one out of six patients and made at least one correct prediction in three patients.

## Discussion

This study provides a critical evaluation of recent deep learning models for detecting FCD in MRI scans from epilepsy patients. Identifying a structural lesion on an MRI plays an important role in the diagnostic process, but is also a strong indicator of subsequent development of drug resistance^23^. For drug-resistant focal epilepsy patients referred for epilepsy surgery evaluation, identifying an epileptogenic lesion on MRI is the most significant predictor of favorable post-surgical outcome^4,24^. However, 15 to 30% of patients with drug-resistant epilepsy are classified as MRI-negative^1^, emphasizing the need for improved tools to detect FCD and other cortical malformations. The main objective of our study was to assess and compare the performance of these classifiers using the assessment from neuroradiology experts as ground truth.

Despite high sensitivities, we find that all classifiers suffer from relatively low PPV. The performance of the DeepFCD tool depended heavily on an empirically determined threshold and achieved at most an accuracy of 56%. Implementing significant threshold variations, or even adaptive thresholding for individual patients, as suggested by the developers^17^, complicates the clinical integration of this tool. MELDGraph outperformed all other classifiers in accuracy and PPV at both the patient- and lesion-level, and showed higher sensitivity than DeepFCD at the 0.9 threshold, marking a significant improvement over its predecessor. However, our results were slightly lower than those reported in the original publication^16^, possibly due to our control group comprising epilepsy patients rather than healthy individuals. Notably, up to 32.8% of MELDGraph’s false positive predictions occurred within regions of non-FCD cortical pathologies, which may hold clinical value, as well as further highlight the challenge of training classifiers specifically to identify FCD.

In a clinical context, an epileptogenic lesion - whether identified through visual inspection or using a classifier - can never stand alone. Any such finding must always be interpreted in the context of seizure semiology, EEG, and potentially other modalities such as FDG-PET, SISCOM-SPECT, MEG, and stereo-EEG^24^. While examining the performance of the classifiers in comparison to neuroradiologists’ assessments, evaluating the potential of these tools to enhance clinical diagnosis falls outside the scope of our study. Work presented by Walger et al. at OHBM 2023^25^ aims to evaluate this interaction between humans and artificial intelligence by integrating the tools into the diagnostic workflow of clinicians. This research is crucial, as it represents the next step in advancing deep learning models towards practical use. Walger et al.^2^ highlight two key scenarios where automatic detection could be particularly relevant: (1) determining the pathological nature of abnormal areas in MRIs and (2) in cases where clinical indicators, such as well-defined regions with continuous EEG spikes or focal FDG-PET hypometabolism, suggest FCD in patients with otherwise ‘MRI-negative’ results. Both application strategies are assessed here: (1) Classifiers tended to misidentify other pathologies as FCD, which could have implications in clinical practice and (2) no significant difference was found between MRI-negative status and classifier sensitivity. The term “MRI-negative” is used inconsistently in the literature. In some cases, it refers to patients whose MRI scans show no visible epileptogenic lesion upon initial review by neuroradiologists^14^. In other cases, it refers more broadly to situations where MRI ultimately fails to contribute to lesion identification, even after re-evaluation^26,2^. In this study, the first interpretation was adopted.

Relatively low specificities underscore the need for incorporating interpretability into neural network outputs. The classifiers aim to promote this by integrating tools such as saliency scores^14^ and confidence. As illustrated in Figure 5A, the DeepFCD and MELDGraph confidence values of TP predictions largely overlap with those of FP predictions. These findings question whether clinicians could reliably distinguish between TP and FP cases when using these tools. Future research should go deeper into methods that effectively contribute to the interpretability of the results.

As indicated here and in a previous study^14^, the quality of preprocessing may influence the performance of the MELD tools. Comprehensive data preprocessing, including manual corrections of tissue segmentation, is necessary to increase the precision of extracted MRI features^27^. This challenge is further compounded by the inclusion of subjects as young as three years old, a practice suggested by the developers of the classifier^14^. FreeSurfer is developed for the segmentation of adult brains from MR images and is unsuitable for children below six years old due to differences in brain morphology and tissue contrast^28^. The integration of the pediatric version of FreeSurfer^29^ into these tools may eventually improve their performance.

Test-retest analysis revealed low levels of consistency in the predictions of all classifiers, indicating that a considerable portion of clusters may be strongly influenced by noise in the images. Future work should aim to understand and account for the sources of these random variations to improve reliability, such as through optimizations in preprocessing and training strategy.

The evaluation was conducted on a dataset that accurately mirrors the varied pathology and image quality found in scans of epilepsy patients. Each subject included in the study had references to focal cortical dysplasia in their MRI reports, affirming its relevance and ensuring that our evaluation context closely resembled clinical settings. This study includes a higher sample size compared to most of the previous independent assessments^14,15,17^ and has the advantage of a balanced cohort based on a single-center dataset of consistent scanner, and sequence parameters compared to previous larger assessments^30^. We employed neuroradiologists’ descriptions and brain atlases, aligning our method with clinical practices, and thereby increasing the relevance and applicability of our findings.

To facilitate data harmonization for the MELD tools, we confined the analysis to a single scanner including T1w and FLAIR MRI, thus substantially reducing the dataset size. In the absence of postoperative scans, the accuracy of the ground truth relied heavily on the precision of the neuroradiologists’ descriptions, with any ambiguity or errors potentially impacting the assessment. Variations in interpretations among radiologists could introduce inconsistencies in the reference standard.

## Conclusion

This study evaluated the performance of state-of-the-art deep learning classifiers for detecting FCD in epilepsy patients. Despite high sensitivities, this study finds that classifiers generally performed with low specificities. Test-retest analysis further revealed inconsistencies, emphasizing the need for more robust models. Future work should focus on improving classifier accuracy, and reliability and integrating interpretability tools. Despite current limitations, these tools may still aid neuroradiologists in detecting subtle FCD cases when combined with multimodal clinical assessments.

## Acknowledgments

This work was supported by the Elsass Foundation and the Lundbeck Foundation (grant R279-2018-1145, BrainDrugs).

## Data Availability

The data used in this study contains personal information and therefore underlies GDPR. Therefore, it cannot be shared openly, but a request to share it securely under a data usage agreement can be made. The code used for this project is openly available at: https://github.com/helenekaas/FCD-tools-evaluation-template.

## Ethical Standards

The work follows appropriate ethical standards in conducting research and writing the manuscript, following all applicable laws and regulations regarding treatment of animals or human subjects.

