## Supplementary Figure 1 for "Independent Evaluation of Deep Learning Models for Detecting Focal Cortical Dysplasia"

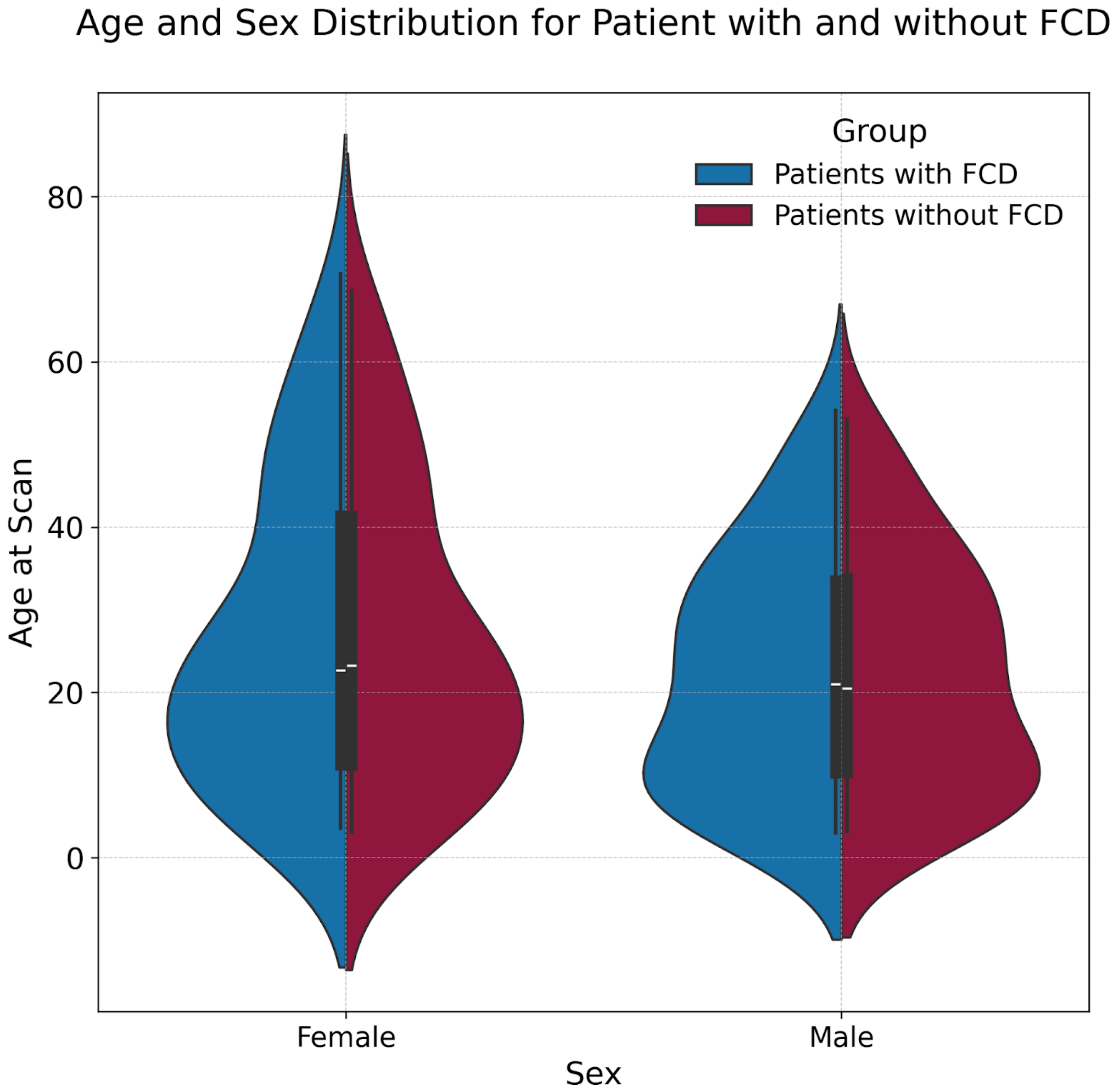


Supplementary Figure 1: The violin plot displays the distribution of age at scan for female and male participants, comparing patients with FCD (blue) to patients without FCD (red).
