## Supplementary Table 1 for "Independent Evaluation of Deep Learning Models for Detecting Focal Cortical Dysplasia"

**Supplementary Table 1:** Classifier Prediction Outputs Sorted According to Ground Truth

| | Patient-level | | | | | Lesion-level | | | | |
|---|---|---|---|---|---|---|---|---|---|---|
| | DeepFCD | | MELD Classifier | MELD Graph | MELD Graph | DeepFCD | | MELD Classifier | MELD Graph | MELD Graph |
| | TH=0.7 | TH=0.90 | - | harmo | no harmo | TH=0.7 | TH=0.9 | - | harmo | no harmo |
| TP | 101 | 63 | 92 | 77 | 79 | 93 | 34 | 107 | 54 | 55 |
| FP | 101 | 51 | 87 | 54 | 69 | 637 | 146 | 709 | 98 | 119 |
| FN | 0 | 38 | 9 | 24 | 22 | - | | - | - | - |
| TN | 0 | 50 | 14 | 47 | 32 | - | | - | - | - |

*TP* True Positive, *FP* False Positive, *TN* True Negative, *FN* False Negative, *TH* Threshold, *harmo* harmonization
