## Supplementary Table 2 for "Independent Evaluation of Deep Learning Models for Detecting Focal Cortical Dysplasia"

**Supplementary Table 2:** Classifier Evaluation Metrics

| **Model** | **Sensitivity** | **Sensitivity CI** | **Specificity** | **Specificity CI** | **Accuracy** | **Accuracy CI** | **PPV** | **PPV CI** | **NPV** | **NPV CI** |
|---|---|---|---|---|---|---|---|---|---|---|
| Patient-level MELD Classifier | 0.91 | 0.85-0.96 | 0.14 | 0.07-0.21 | 0.52 | 0.46-0.59 | 0.51 | 0.44-0.59 | 0.61 | 0.43-0.83 |
| Patient-level MELDGraph (harmo) | 0.76 | 0.67-0.84 | 0.47 | 0.38-0.56 | 0.61 | 0.55-0.68 | 0.59 | 0.50-0.67 | 0.66 | 0.55-0.77 |
| Patient-level MELDGraph (no harmo) | 0.78 | 0.70-0.86 | 0.32 | 0.23-0.42 | 0.55 | 0.48-0.62 | 0.53 | 0.45-0.61 | 0.59 | 0.46-0.70 |
| Patient-level DeepFCD (TH=0.7) | 1 | 1.0-1.0 | 0 | 0.0-0.0 | 0.5 | 0.44-0.56 | 0.5 | 0.43-0.57 | 0 | 0-0 |
| Patient-level DeepFCD (TH=0.9) | 0.62 | 0.53-0.71 | 0.5 | 0.40-0.59 | 0.56 | 0.49-0.62 | 0.55 | 0.46-0.64 | 0.57 | 0.47-0.67 |
| Lesion-level MELDClassifier | 0.92 | 0.87-0.97 | - | - | - | - | 0.13 | 0.11-0.16 | - | - |
| Lesion-level MELDGraph (harmo) | 0.69 | 0.59-0.79 | - | - | - | - | 0.36 | 0.28-0.43 | - | - |
| Lesion-level MELDGraph (not harmo) | 0.71 | 0.61-0.82 | - | - | - | - | 0.32 | 0.25-0.38 | - | - |
| Lesion-level DeepFCD (TH=0.7) | 1 | 1.0-1.0 | - | - | - | - | 0.04 | 0.03-0.04 | - | - |
| Lesion-level DeepFCD (TH=0.9) | 0.47 | 0.35-0.68 | - | - | - | - | 0.19 | 0.13-0.25 | - | - |

*PPV* Positive Predictive Value Positive, *NPV* Negative Predictive Value, *TH* Threshold, *harmo* harmonization, *CI* Confidence Interval
