## Supplementary Table 3 for "Independent Evaluation of Deep Learning Models for Detecting Focal Cortical Dysplasia"

**Supplementary Table 3:** Overview of Other Pathologies and Detection Rate per Classifier

| **Unique Value** | **Count** | DeepFCD | MELD Classifier | MELD Graph (harmo) | MELD Graph (no harmo) | DeepFCD Mean Confidence | DeepFCD Mean Probability | MELD Graph (harmo) Mean Probability | MELD Graph (no harmo) Mean Probability |
|---|---|---|---|---|---|---|---|---|---|
| **Incomplete inversion of the hippocampus** | 74 | 64 | 15 | 4 | 8 | 63.482258 | 0.804507 | 15.293333 | 30.148571 |
| **Cyst** | 14 | 8 | 6 | 2 | 4 | 58.142857 | 0.792546 | 18.27 | 16.23 |
| **MTS** | 13 | 11 | 4 | 6 | 7 | 81.209091 | 0.833422 | 29.056667 | 33.997143 |
| **DVA** | 11 | 8 | 4 | 1 | 3 | 53.35 | 0.79267 | 14.15 | 18.03 |
| **Lack of myelination** | 8 | 8 | 3 | 2 | 3 | 48.5625 | 0.748725 | 17.205 | 23.525 |
| **Folding abnormalities** | 7 | 5 | 4 | 0 | 3 | 62.96 | 0.808851 | - | 42.063333 |
| **Trauma sequelae** | 6 | 4 | 4 | 2 | 2 | 90 | 0.847388 | 43.855 | 53.265 |
| **Infarction** | 5 | 4 | 4 | 4 | 4 | 85.95 | 0.835867 | 65.075 | 65.9425 |
| **Subcortical WML** | 5 | 0 | 0 | 0 | 0 | - | - | - | - |
| **Ganglioglioma** | 5 | 5 | 2 | 2 | 0 | 79.64 | 0.824165 | 40.855 | - |
| **Heterotopia** | 4 | 2 | 2 | 1 | 1 | 56.1 | 0.806644 | 73.74 | 69.06 |
| **Tumor, unspecified** | 2 | 2 | 1 | 1 | 0 | 95.8 | 0.859887 | 13.75 | - |
| **Falx calcification** | 2 | 1 | 0 | 0 | 0 | 47.7 | 0.776561 | - | - |
| **Gliosis** | 2 | 1 | 0 | 0 | 1 | 95.8 | 0.859887 | - | 14.8 |
| **Hemorrhage** | 1 | 1 | 1 | 0 | 0 | 99.8 | 0.872652 | - | - |
| **Polymicrogyria** | 1 | 1 | 1 | 0 | 0 | 56.1 | 0.806644 | - | - |
| **Aneurysm** | 1 | 0 | 1 | 0 | 0 | - | - | - | - |
| **DNET** | 1 | 1 | 0 | 0 | 1 | 52.2 | 0.77653 | - | 6.98 |
| **Hemangioma** | 1 | 1 | 1 | 0 | 0 | 95.8 | 0.859887 | - | - |
| **SWS** | 1 | 1 | 1 | 1 | 1 | 99.8 | 0.872652 | 26.84 | 21.45 |
| **Migration abnormalities** | 1 | 1 | 0 | 1 | 1 | 45.3 | 0.790605 | 92.16 | 92.98 |
| **Encephalocele** | 1 | 0 | 0 | 0 | 0 | - | - | - | - |
| **Perinatal ischemia** | 1 | 1 | 1 | 0 | 0 | 99.8 | 0.872652 | - | - |

*DNET* Dysembryoplastic Neuroepithelial Tumor, *SWS* Sturge-Weber Syndrome, *MTS* Mesial Temporal Sclerosis, *DVA* Developmental Venous Anomaly, WML White Matter Lesions
