## Supplementary Text 1 for "Independent Evaluation of Deep Learning Models for Detecting Focal Cortical Dysplasia"

**Supplementary Text 1: MRI Data**

The most frequent T1w and FLAIR image from the data extraction had the sequence names "“t1_mpr_sag_iso_neuronav” and “t2_spc_da-fl_irprep_sag_p2_iso”, respectively. For all patients, the parameters for the T1w Magnetization Prepared Rapid Gradient Echo were: repetition time = 1900 ms, echo time [ET] = 2.23 ms, flip angle = 9°, slice thickness = 1 mm, voxel size = 1mm x 1mm x 1mm. The parameters for the FLAIR sequence were: repetition time = 5000 ms, echo time = 395 ms, flip angle = 12°, slice thickness = 1 mm, voxel size = 1mm x 1mm x 1mm].
