## Supplementary Figure 2 for "Independent Evaluation of Deep Learning Models for Detecting Focal Cortical Dysplasia"

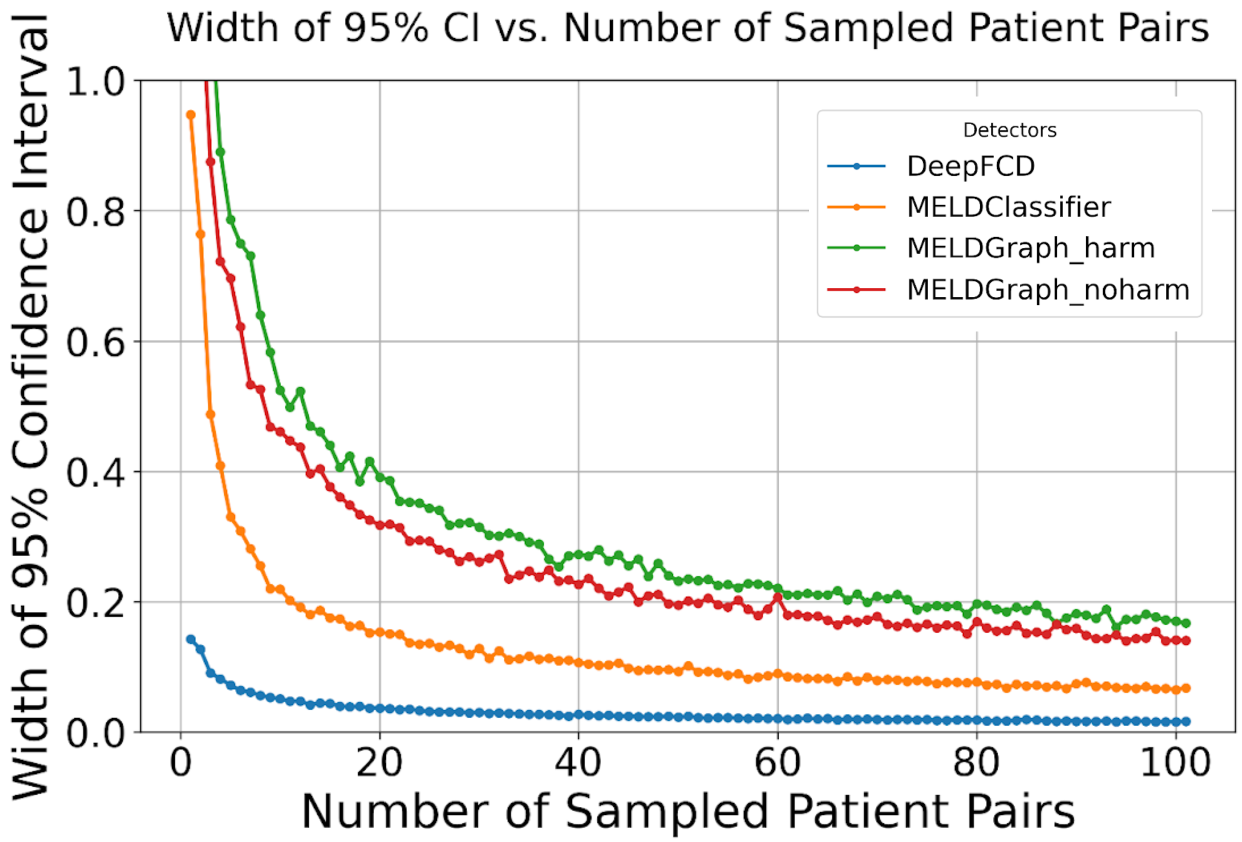


Supplementary Figure 2: Bootstrapped accuracy showing the width of the 95% confidence intervals as a function of sampled patient pairs. Accuracy was estimated on matched patient pairs using 1,000 bootstrap iterations per sample size. The x-axis represents the number of sampled patient pairs, and the y-axis shows the width of the confidence interval for estimated accuracy.
